# Multisite Pragmatic Cluster-Randomized Controlled Trial of the CONCERN Early Warning System

**DOI:** 10.1101/2024.06.04.24308436

**Authors:** Sarah C. Rossetti, Patricia C. Dykes, Chris Knaplund, Sandy Cho, Jennifer Withall, Graham Lowenthal, David Albers, Rachel Lee, Haomiao Jia, Suzanne Bakken, Min-Jeoung Kang, Frank Y. Chang, Li Zhou, David W. Bates, Temiloluwa Daramola, Fang Liu, Jessica Schwartz-Dillard, Mai Tran, Syed Mohtashim Abbas Bokhari, Jennifer Thate, Kenrick D. Cato

## Abstract

**Importance:** Late predictions of hospitalized patient deterioration, resulting from early warning systems (EWS) with limited data sources and/or a care team’s lack of shared situational awareness, contribute to delays in clinical interventions. The COmmunicating Narrative Concerns Entered by RNs (CONCERN) Early Warning System (EWS) uses real-time nursing surveillance documentation patterns in its machine learning algorithm to identify patients’ deterioration risk up to 42 hours earlier than other EWSs.

**Objective:** To test our a priori hypothesis that patients with care teams informed by the CONCERN EWS intervention have a lower mortality rate and shorter length of stay (LOS) than the patients with teams not informed by CONCERN EWS.

**Design:** One-year multisite, pragmatic controlled clinical trial with cluster-randomization of acute and intensive care units to intervention or usual-care groups.

**Setting:** Two large U.S. health systems.

**Participants:** Adult patients admitted to acute and intensive care units, excluding those on hospice/palliative/comfort care, or with Do Not Resuscitate/Do Not Intubate orders.

**Intervention:** The CONCERN EWS intervention calculates patient deterioration risk based on nurses’ concern levels measured by surveillance documentation patterns, and it displays the categorical risk score (low, increased, high) in the electronic health record (EHR) for care team members.

**Main Outcomes and Measures:** Primary outcomes: in-hospital mortality, LOS; survival analysis was used. Secondary outcomes: cardiopulmonary arrest, sepsis, unanticipated ICU transfers, 30-day hospital readmission.

**Results:** A total of 60 893 hospital encounters (33 024 intervention and 27 869 usual-care) were included. Both groups had similar patient age, race, ethnicity, and illness severity distributions. Patients in the intervention group had a 35.6% decreased risk of death (adjusted hazard ratio [HR], 0.644; 95% confidence interval [CI], 0.532-0.778; P<.0001), 11.2% decreased LOS (adjusted incidence rate ratio, 0.914; 95% CI, 0.902-0.926; P<.0001), 7.5% decreased risk of sepsis (adjusted HR, 0.925; 95% CI, 0.861-0.993; P=.0317), and 24.9% increased risk of unanticipated ICU transfer (adjusted HR, 1.249; 95% CI, 1.093-1.426; P=.0011) compared with patients in the usual-care group.

**Conclusions and Relevance:** A hospital-wide EWS based on nursing surveillance patterns decreased in-hospital mortality, sepsis, and LOS when integrated into the care team’s EHR workflow.

**Trial Registration:** ClinicalTrials.gov Identifier: NCT03911687 https://clinicaltrials.gov/ct2/show/NCT03911687

**Key Points:** *Question:* Do patients whose care team receive the CONCERN Early Warning System (EWS) intervention have a lower mortality rate and shorter length of stay than patients in the usual-care group?

*Findings:* In this multisite, pragmatic cluster-randomized controlled clinical trial that included 60 893 hospital patient encounters, patients whose care team received the CONCERN EWS intervention had a 35.6% decreased risk of death and 11.2% shorter length of stay compared with those in the usual-care group.

*Meaning:* A machine learning-based EWS modeled on nursing surveillance patterns significantly decreased the risk of inpatient deterioration events.

## 1. Introduction

Given the rise in patient acuity^1^, early identification of patients’ risk of deterioration is essential to preventing avoidable yet serious adverse hospital outcomes^2^, such as mortality and sepsis. Failure to detect deterioration and intervene accordingly is an unacceptable system failure, strongly linked to information and communication breakdowns among the care team.^3^ While several non-randomized studies have shown that automated algorithm-based Early Warning Systems (EWSs) positively impact patient outcomes, few randomized controlled trials have demonstrated an impac^4,5^ and many predictions are focused on one particular event type rather than a broad set of outcomes, such as in-hospital mortality, length of stay (LOS) and sepsis.^4,6^ Advances in EWS computational sophistication hold great promise for predicting patient “crashing”^7^, however algorithms typically rely on late and noisy physiologic indicators of deterioration (eg, labs, vital signs).^8,9^ Novel approaches are needed to both identify deterioration earlier and make the care team aware so that timely interventions can be performed.

Nurse surveillance is a core component of nursing practice aimed at preventing adverse events.^10^ Increased nurse surveillance is an indicator of concern^11,12^, and nurse concern has been shown to be a valid and frequent reason for calling a rapid response.^12^ Nurses can recognize subtle, yet observable, clinical changes that may not be captured in physiological data or well-displayed in electronic health records (EHRs)^11^, such as pallor change with incremental increases in supplemental oxygen needs, slower recovery of arterial blood pressure after turning the patient, or small changes in mental status from baseline. As part of surveillance, nurses document additional data in the EHR to highlight concerning patient changes, but the patterns of these additional data are not explicitly evident to other members of the care team.^11,13^ Unfortunately, when there is a lack of shared team situational awareness, medical interventions are delayed.^11–14^

Evidence for escalation of medical interventions based on nurses’ concerns has long remained classified as level 5 evidence (expert opinion).^15–20^ To objectively measure and test nurses’ concern levels in predicting patient deterioration, we created a machine learning-based predictive model—the COmmunicating Narrative Concerns Entered by RNs (CONCERN) EWS—that processes nurse surveillance patterns from structured and narrative documentation.^12^ The model continuously monitors nurses’ concern levels, assigns a categorical deterioration risk score of green (low), yellow (increased), or red (high), updates the score hourly, and the CONCERN clinical-decision-support EWS displays the score in the EHR for care team members. CONCERN EWS research findings have demonstrated that it can predict patient deterioration 42 hours earlier than other leading EWSs^12^, and nurses and prescribing providers perceive situational awareness is enhanced through its use.^21^ Earlier predictions provide clinicians with greater lead time for action^22^, ultimately enabling the care team to identify patients who may be *entering* a risky state with enough time to intervene.^12^ Therefore, the objective of this study was to determine at the individual patient level whether CONCERN EWS led to a decrease in primary outcomes (in-hospital mortality and LOS) and influenced secondary outcomes (cardiopulmonary arrest, sepsis, unanticipated ICU transfers, and 30-day hospital readmission). We hypothesized a priori that patients whose care team received CONCERN EWS risk scores would have lower mortality and LOS than a control group of patients with non-EWS-informed teams.

## 2. Methods

### 2.1 Study Design, Trial Sites, and Randomization

We evaluated CONCERN EWS in a 1-year multisite, pragmatic, cluster-randomized controlled clinical trial at 2 large health systems in the Northeastern United States. Each system was a study site (A and B), and sites A and B each comprise 1 academic medical center and 1 community hospital. Individual study units across the 4 hospitals included all non-specialty acute care units (ACUs) and ICUs. Specialty ACUs, such as oncology, psychiatric, and rehabilitation, were excluded. Randomization was performed at the unit level and stratified by site prior to trial initiation.^23^ Each study unit (ACU or ICU) was randomly allocated to one of 2 groups (CONCERN EWS intervention or usual-care) using a computer-generated randomization scheme.^23^ The trial included 74 clinical units (37 intervention, 37 usual-care) with the following distribution: Site A intervention - 19 ACUs and 5 ICUs; Site A usual-care - 17 ACUs and 7 ICUs; Site B intervention - 9 ACUs and 4 ICUs; Site B usual-care - 8 ACUs and 5 ICUs (eTable1). The intervention (CONCERN EWS prediction risk score) was displayed to a patient’s care team consisting of nurses and prescribing providers (i.e., physicians, nurse practitioners, physician assistants) only if that patient was admitted to an intervention-assigned unit. The intervention was not displayed to care teams of patients admitted to usual-care-assigned units.

Due to unavoidable delays related to the COVID-19 pandemic, the study time frame differed for each site: the trial was conducted at Site A from October 2020 to October 2021 and at Site B from October 2021 to October 2022. The original protocol, published previously, described multiple time-series intervention with non-equivalent control groups.^24^ But prior to beginning the study, we determined cluster-randomization of study units was feasible and modified the protocol. See eMethods1 for modifications and rationale. Institutional review boards (IRBs) at each site approved the protocol before trial initiation with a waiver of consent.

### 2.2 Trial Participants and Outcomes

While randomization occurred at the unit level, outcomes were assessed at the patient level due to patient movement across units during their hospitalization. Patients on study units were included in analyses if they were 18 years of age and older, hospitalized for greater than 24 hours (EWS score begins displaying after 24 hours), admitted to a study unit for a minimum of 12 hours, and free from any in-hospital event (including discharge) until at least 6 hours after study unit admission. Hospice and palliative care patients and patients with do not resuscitate/do not intubate and comfort care orders activated prior to any trial outcome event were excluded.

Primary outcomes were in-hospital mortality rate and LOS. Secondary outcomes were rates of cardiopulmonary arrest, sepsis^25^, unanticipated ICU transfer, and 30-day hospital readmission.^26^ See Table 1 for definitions. All data were collected from the EHR and evaluated and reported across study sites.

**Table 1.** Definitions of Study Outcomes.

| Study Outcome |  | Definition |
| --- | --- | --- |
| Primary | In-hospital mortality | Patients who died while in the hospital. |
|  | Length of Stay (LOS) | Calculated time elapsed between patient admission to an inpatient care unit and hospital discharge, separated from the hospital, or died during hospitalization. LOS calculations excluded pre-inpatient hospital time, such as emergency department time. |
| Secondary | Cardiopulmonary arrest | A Code Blue event that occurred while hospitalized on a study unit for at least 12 hours. |
|  | Sepsis | Sepsis-3<sup>25</sup> guideline of concurrent suspected infection-positive (body fluid cultures ordered, and oral or parenteral antibiotics administered within a specified time range) and Sequential Organ Failure Assessment (SOFA) or quick (q)SOFA positive criteria within specified time windows of one another. |
|  | Unanticipated ICU transfer | Transfer from a 12-hour or longer stay on an ACU directly to the ICU, not resulting from a planned procedure. |
|  | 30-day hospital readmission | Unplanned readmission of at least 24 hours (for multiple events the earliest is counted) for any cause to an acute care hospital (trackable in the same health system) within 30 days of discharge, excluding planned, same-day, and other specific types of readmissions.<sup>26</sup> |

### 2.3 CONCERN EWS Intervention

CONCERN EWS comprises a predictive model that uses an ensemble^27^ machine learning approach to process nursing surveillance patterns in the EHR, predicts clinical deterioration in the next 24 hours, and displays the CONCERN EWS risk score as a green, yellow, or red icon (non-interruptive alert) on the EHR patient list (main landing screen upon login) (Figure 1). The icon is visible to every nurse and prescribing provider on the patient’s clinical team, and the team also can double-click the icon to access a screen with prediction details. By targeting the entire clinical team rather than a solitary provider, CONCERN EWS promotes shared situational awareness. All existing hospital policies and procedures related to patient deterioration and escalation of care remained the same across intervention and usual-care groups.

**Figure 1.**
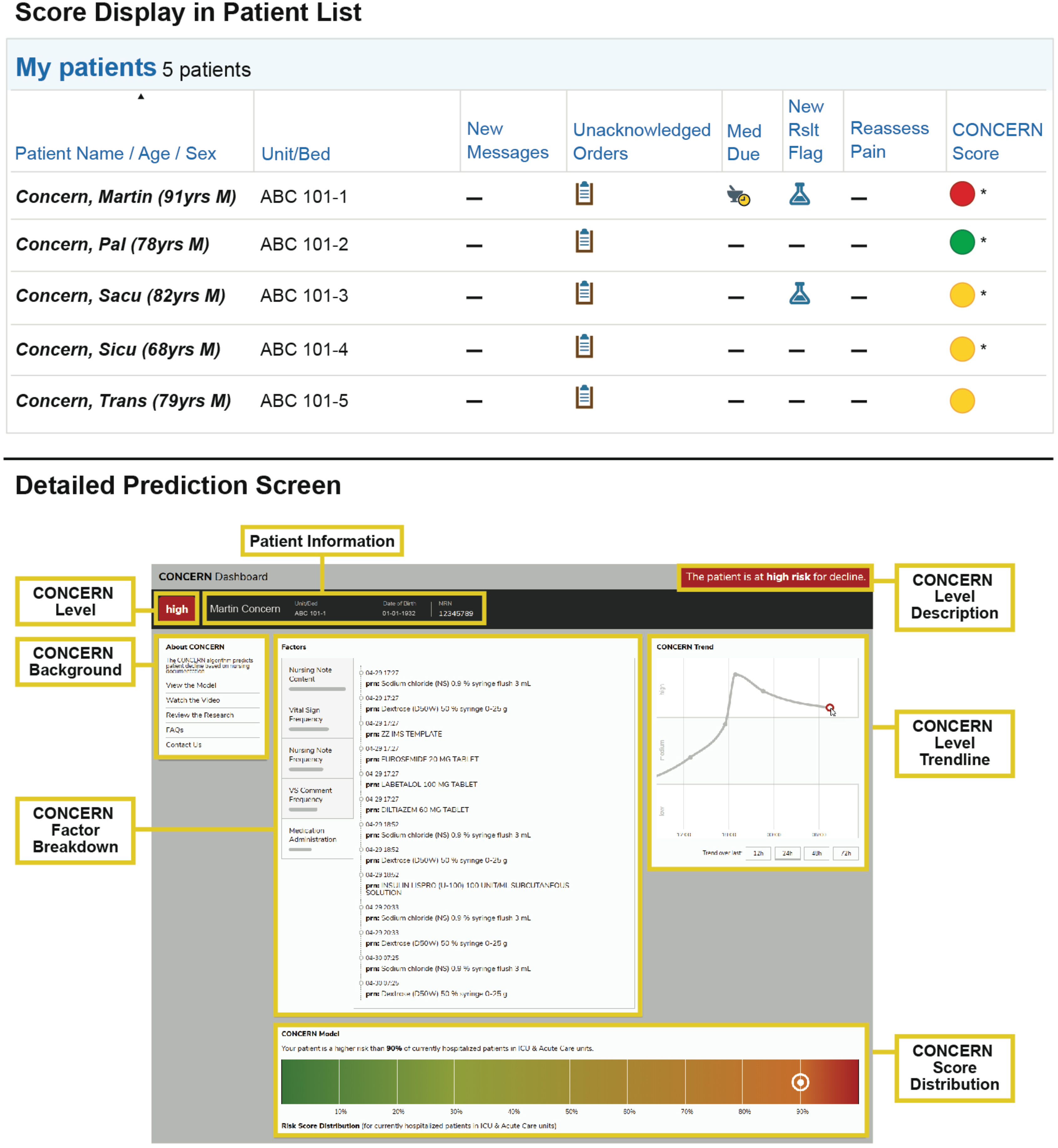
CONCERN EWS Display and Detailed Prediction Screen integrated into the EHR.

Details of CONCERN EWS modeling approach (eFigure1), performance (eTable 2), factors and features (eTable3), and EHR integration (eMethods2) have been published elsewhere.^12,21,28^ Briefly, model development used 217,166 distinct inpatient encounters between 2015 and 2019 from 2 study sites, with a task of predicting a deterioration event in the next 24 hours based on the patient’s preceding 24-hour data. During model development, the first occurrence of cardiopulmonary arrest, rapid response, sepsis, transfer to ICU and death were identified as proxy measures for patient deterioration. The gradient-boosted decision-tree model was trained at site B on 70% of a retrospective dataset, with 30% used for 10-fold cross-validation, with greater than 97% average accuracy and 95% average area under the curve (eTable2, eMethods2) and was externally validated at site A.

### 2.4 Statistical Analysis

The primary objective was to test the hypothesis that CONCERN EWS decreases in-hospital mortality and LOS (independent outcomes). Secondary outcomes were CONCERN EWS’s influence on in-hospital events (ie, sepsis, cardiopulmonary arrest, unanticipated ICU transfers) and 30-day hospital readmissions. Power analysis comparing mortality rates between groups estimated a 1-year trial on the targeted units at our 2 sites would result in sufficient sample size for at least 80% statistical power to detect a difference of less than 1% relative difference in mortality rates (2-sided; α=.05) (eTable4).

We originally planned to analyze at the unit level, but because patients moved across different units all outcomes were analyzed at the individual patient level per hospital encounter (multiple hospital encounters possible) (eMethods1). All regression models (generalized linear model (GLM) and Cox proportional hazard (PH) models) included the following covariates: Charlson Comorbidity Index, age, sex, race, ethnicity, and study site. In addition to these covariates, PH models also included ICU as a time dependent covariate. All analyses accounted for clustering at the unit level.

#### 2.4.1 LOS Outcome

We compared LOS between intervention and usual-care by: 1) survival analysis using PH model, and 2) GLM analysis. PH model with time-dependent explanatory variables tested the hazard ratio for time-to-discharge (censored at death) between the 2 groups. The intervention group was a time-dependent explanatory variable. PH model used a marginal frailty model. For GLM analysis, LOS was calculated as the time from a patient’s admission to an inpatient care unit to discharge or death (if died).^29^ GLM tested incident rate ratio for LOS between the 2 groups with log-link and negative binomial distribution.^29^ We estimated mean LOS from GLM. GLM used sandwich standard error estimates.

#### 2.4.2 Death and Secondary In-Hospital Event Outcomes

We used survival analysis to compare time-to-hospital-event outcomes between groups.^30^ We examined time-to-death and time-to-each of the secondary outcome in-hospital events (sepsis, cardiopulmonary arrest, unanticipated ICU transfers) using PH model with time-dependent explanatory variables. The time-to-event analysis stops at the first outcome observed. If patients died, had any of the other two in-hospital events, or were discharged before the first occurrence of the in-hospital event of interest, time to the event outcome was censored at death, the first occurrence of the other in-hospital events, or discharge. Event timing was measured in 12-hour increments to align with the duration of typical clinical shifts. Table 3 provides event definitions.

#### 2.4.3 Transfers Between Intervention- and Usual-Care Units

Hospitalized patients are routinely transferred between units due to changing care needs. In this pragmatic trial, we employed several methods to control for patients who were transferred between units randomized as intervention and usual-care.^23^ Survival analysis defined the event location as the unit the patient was on for the majority of the time during the 12-hour shift when an event outcome occurred. GLM analysis for LOS labeled the encounter as either intervention or usual-care group according to which unit the patient spent most time during the hospitalization. We were unable to draw Kaplan-Meier survival curves by group because: 1) intervention status is a time dependent variable and 2) ICU status, a covariate, also is a time-dependent variable.

No outcomes were measured across a patient’s multiple hospital encounters. A 30-day hospital readmission was treated as an outcome of the original hospital encounter not a new hospital encounter. We calculated the odds ratio for 30-day hospital readmission using logistic generalized linear mixed model.

## 3. Results

### 3.1 Trial Participants and Hospital Encounters

During the study timeframes at sites A and B, 58 994 unique patients (79 049 hospital encounters) were admitted to study units. After excluding 18 156 encounters that did not meet eligibility criteria, 60 893 encounters were included in trial analyses, with a total of 33 024 encounters in the CONCERN EWS intervention group and 27 869 in the usual-care group for our survival analyses (Figure 2).

**Figure 2.**
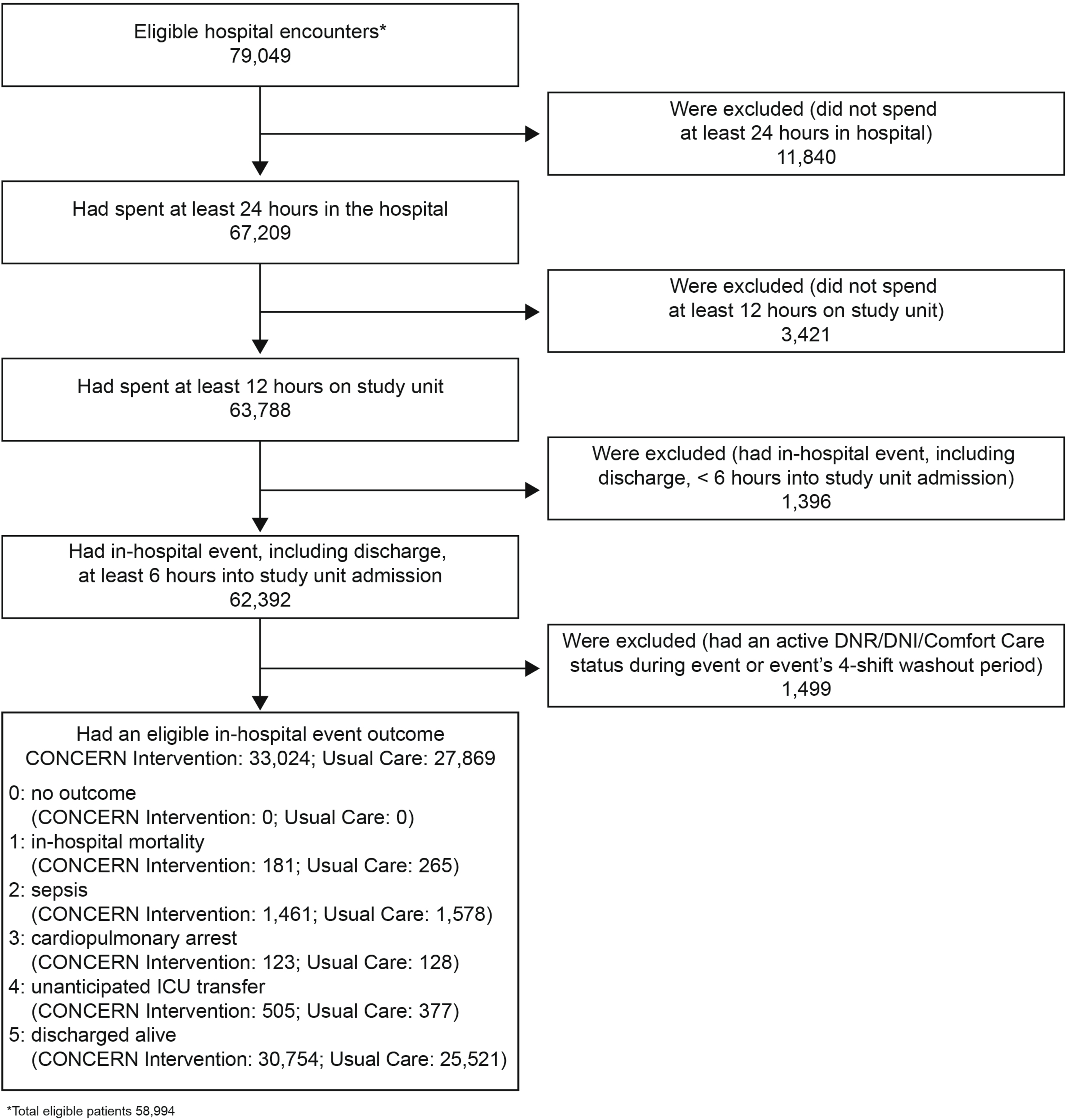
Flow Diagram of Patient Encounters Assessed for Eligibility and Allocation to Intervention or Usual-Care for In-Hospital Event Outcomes.

Overall, patients across intervention and usual-care groups represented similar age, race, ethnicity, and illness severity distributions (Table 2). Illness severity is demonstrated in Table 2 using 2 proxy variables: 1) the Charlson Comorbidity Index and 2) discharge disposition. Additional site comparisons are provided in eTables 5 and 6.

**Table 2.** Characteristics of Patients with Hospital Encounters During the Trial (N=60,893)

|  | CONCERN<br>Intervention<br>(n=33,024<sup>a</sup>) | Usual-Care<br>(n=27,869<sup>a</sup>) | Standardized mean<br>difference<sup>b</sup> |
| --- | --- | --- | --- |
| Age | 62.62 ± 17.56 | 63.67 ± 17.10 | 0.06 |
| Male Sex – no. (%) | 16,056 (48.62) | 13,966 (50.11) | 0.03 |
| Race<sup>c</sup> – no. (%) |  |  |  |
| White | 18,923 (57.30) | 16,278 (58.41) | 0.02 |
| Black | 4,878 (14.77) | 4,224 (15.16) | 0.01 |
| Asian | 703 (2.13) | 634 (2.27) | 0.01 |
| Other or missing | 8,520 (25.80) | 6,733 (24.16) | 0.04 |
| Ethnic group<sup>c</sup> – no. (%) |  |  |  |
| Not Hispanic or Latino | 23,618 (71.52) | 20,304 (72.86) | 0.03 |
| Hispanic or Latino | 7,442 (22.54) | 5,903 (21.18) | 0.03 |
| Unknown or not reported | 1,964 (5.95) | 1,662 (5.96) | 0.0004 |
| Primary Language English – no. (%) | 26,713 (80.89) | 22,584 (81.04) | 0.004 |
| Charlson comorbidity | 3.11 ± 3.16 | 3.51 ± 3.34 | 0.12 |
| Discharge Disposition<sup>d</sup> – no. (%) |  |  |  |
| Home | 26,773 (81.07) | 22,206 (79.68) | 0.04 |
| Other | 6,251 (18.93) | 5,663 (20.32) | 0.04 |
| <sup>a</sup>Reported N for intervention and usual-care refers to survival analysis.<br><sup>b</sup>All standardized mean differences are <0.2, therefore no between group differences<br><sup>c</sup>Race and ethnic group were reported in the Electronic Health Record<br><sup>d</sup>Discharge Disposition: Home includes home with services; Other includes any disposition not to home<br>± Plus-minus values are mean ±SD |  |  |  |

### 3.2 Primary Outcomes

The intervention group had a lower risk of in-hospital mortality and decreased LOS. A total of 181 patients in the intervention and 265 patients in usual-care experienced in-hospital mortality as the first negative event in our survival analysis, indicating a 35.6% decreased instantaneous risk of dying in the hospital with the CONCERN intervention (adjusted hazard ratio [HR], 0.644; 95% confidence interval [CI], 0.532-0.778; P<.0001) (Table 3).

**Table 3.**
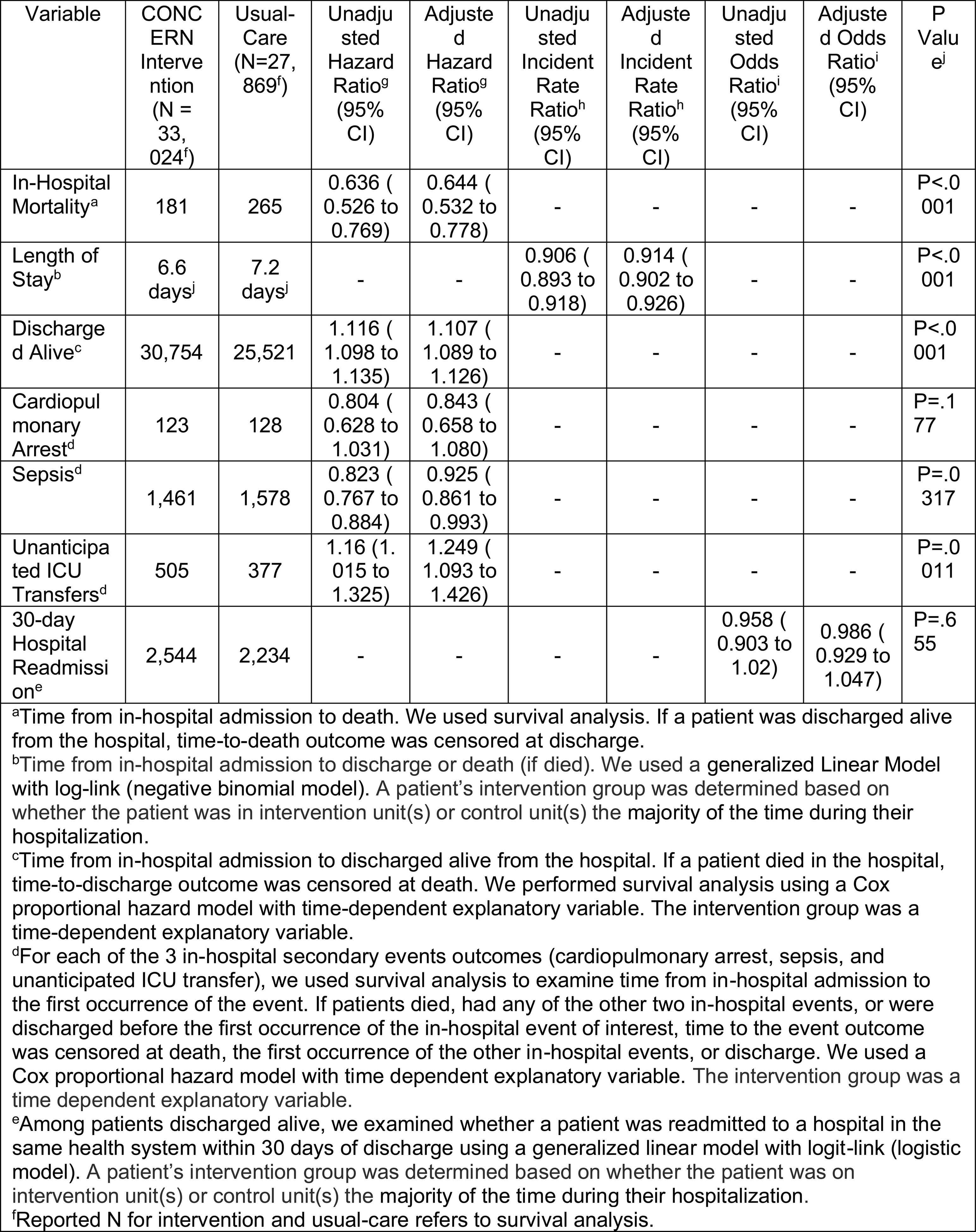
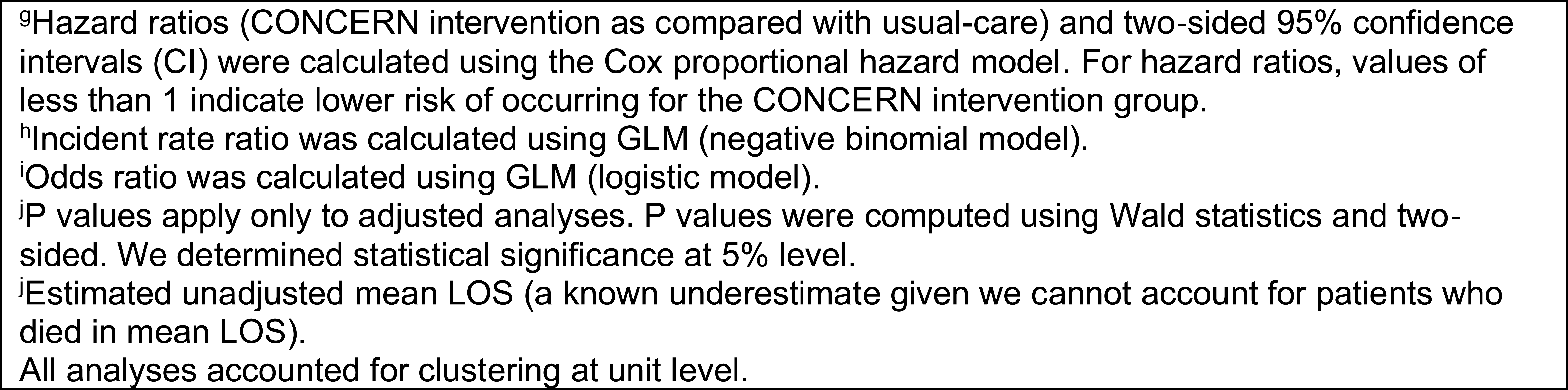
CONCERN Impact on Primary and Secondary Outcomes.

| Variable | CONCERN Intervention (N = 33,024<sup>f</sup>) | Usual-Care (N=27,869<sup>f</sup>) | Unadjusted Hazard Ratio<sup>g</sup> (95% CI) | Adjusted Hazard Ratio<sup>g</sup> (95% CI) | Unadjusted Incident Rate Ratio<sup>h</sup> (95% CI) | Adjusted Incident Rate Ratio<sup>h</sup> (95% CI) | Unadjusted Odds Ratio<sup>i</sup> (95% CI) | Adjusted Odds Ratio<sup>i</sup> (95% CI) | P Value<sup>j</sup> |
| --- | --- | --- | --- | --- | --- | --- | --- | --- | --- |
| In-Hospital Mortality<sup>a</sup> | 181 | 265 | 0.636 (0.526 to 0.769) | 0.644 (0.532 to 0.778) | - | - | - | - | P<.0001 |
| Length of Stay<sup>b</sup> | 6.6 days<sup>j</sup> | 7.2 days<sup>j</sup> | - | - | 0.906 (0.893 to 0.918) | 0.914 (0.902 to 0.926) | - | - | P<.0001 |
| Discharged Alive<sup>c</sup> | 30,754 | 25,521 | 1.116 (1.098 to 1.135) | 1.107 (1.089 to 1.126) | - | - | - | - | P<.0001 |
| Cardiopulmonary Arrest<sup>d</sup> | 123 | 128 | 0.804 (0.628 to 1.031) | 0.843 (0.658 to 1.080) | - | - | - | - | P=.177 |
| Sepsis<sup>d</sup> | 1,461 | 1,578 | 0.823 (0.767 to 0.884) | 0.925 (0.861 to 0.993) | - | - | - | - | P=.0317 |
| Unanticipated ICU Transfers<sup>d</sup> | 505 | 377 | 1.16 (1.015 to 1.325) | 1.249 (1.093 to 1.426) | - | - | - | - | P=.0011 |
| 30-day Hospital Readmission<sup>e</sup> | 2,544 | 2,234 | - | - | - | - | 0.958 (0.903 to 1.02) | 0.986 (0.929 to 1.047) | P=.655 |

<sup>a</sup>Time from in-hospital admission to death. We used survival analysis. If a patient was discharged alive from the hospital, time-to-death outcome was censored at discharge.
<sup>b</sup>Time from in-hospital admission to discharge or death (if died). We used a generalized Linear Model with log-link (negative binomial model). A patient's intervention group was determined based on whether the patient was in intervention unit(s) or control unit(s) the majority of the time during their hospitalization.
<sup>c</sup>Time from in-hospital admission to discharged alive from the hospital. If a patient died in the hospital, time-to-discharge outcome was censored at death. We performed survival analysis using a Cox proportional hazard model with time-dependent explanatory variable. The intervention group was a time-dependent explanatory variable.
<sup>d</sup>For each of the 3 in-hospital secondary events outcomes (cardiopulmonary arrest, sepsis, and unanticipated ICU transfer), we used survival analysis to examine time from in-hospital admission to the first occurrence of the event. If patients died, had any of the other two in-hospital events, or were discharged before the first occurrence of the in-hospital event of interest, time to the event outcome was censored at death, the first occurrence of the other in-hospital events, or discharge. We used a Cox proportional hazard model with time dependent explanatory variable. The intervention group was a time dependent explanatory variable.
<sup>e</sup>Among patients discharged alive, we examined whether a patient was readmitted to a hospital in the same health system within 30 days of discharge using a generalized linear model with logit-link (logistic model). A patient's intervention group was determined based on whether the patient was on intervention unit(s) or control unit(s) the majority of the time during their hospitalization.
<sup>f</sup>Reported N for intervention and usual-care refers to survival analysis.
<sup>g</sup>Hazard ratios (CONCERN intervention as compared with usual-care) and two-sided 95% confidence intervals (CI) were calculated using the Cox proportional hazard model. For hazard ratios, values of less than 1 indicate lower risk of occurring for the CONCERN intervention group.
<sup>h</sup>Incident rate ratio was calculated using GLM (negative binomial model).
<sup>i</sup>Odds ratio was calculated using GLM (logistic model).
<sup>j</sup>P values apply only to adjusted analyses. P values were computed using Wald statistics and two-sided. We determined statistical significance at 5% level.
All analyses accounted for clustering at unit level.

There was a 11.2% decrease in mean LOS with the CONCERN intervention (adjusted incidence rate ratio, 0.914; 95% CI, 0.902-0.926; P<.0001). The estimated unadjusted mean LOS (a known underestimate given we cannot account for patients who died) was 157.9 hours (6.6 days) for patients in the intervention and 172.7 hours (7.2 days) for patients in usual-care (Table 3).

### 3.3 Secondary Outcomes

Analyses performed between groups for each outcome identified through our survival analysis as a patient’s first in-hospital event demonstrated that patients in the intervention group had a lower risk of sepsis and a higher risk of unanticipated ICU transfer. Among patients experiencing sepsis as the first hospital event, there were 1,461 patients in the intervention group and 1,578 patients in usual-care group (adjusted HR, 0.925; 95% CI, 0.861-0.993; P=.0317). For unanticipated ICU transfers, there were 505 patients in the intervention group and 377 patients in usual-care group (adjusted HR, 1.249; 95% CI, 1.093-1.426; P=.0011). No statistically significant differences between groups were found for cardiopulmonary arrest (P=.177) or 30-day hospital readmission (P=.655) (Table 3).

## 4. Discussion

Patients whose care teams were informed by CONCERN EWS were a third less likely to die, and a quarter more likely to be transferred to intensive care. Nurses can observe subtle changes which suggest that patients are more likely to deteriorate, and early recognition and treatment of these patients can improve outcomes.

Other EWSs have influenced in-hospital mortality, sepsis, or LOS, but not across the breadth of outcomes we observed.^4,5,31,32^ CONCERN EWS is a single hospital-wide intervention (ie, implemented across ACUs and ICUs) for all-cause deterioration. Most published studies target a particular condition (e.g., sepsis) or hospital setting (e.g., ICU)^4^ and only a minority of EWS have been evaluated in randomized controlled trials.^4,6^ Additionally, a 2022 systematic review identified only 41 randomized controlled trials for machine learning interventions in health care, and none adhered to all CONSORT-AI standards.^33^ We report our trial findings according to the CONSORT guidelines^34^ and extensions for AI^35^, cluster-randomized trials^36^, and pragmatic trials^37^ (eTable7).

Unanticipated ICU transfer increased in the intervention group; early ICU transfer has been shown to be a “window of critical opportunity” for timely clinical interventions that alter the trajectory of a patient’s clinical progression, prevent adverse outcomes, and improve survival.^38–42^ Several methodological choices allowed CONCERN EWS to have accurate and robust predictive power and sufficient lead time to alter a patient’s clinical trajectory, including: 1) robust modeling of temporal data patterns^43–45^ (e.g., time of day, day of week, patient hospital day) for health care processes (e.g., nurses’ surveillance and medication administration decisions) that reflect a nurse’s level of concern about a patient‘s deterioration risk, and 2) the use of nursing assessment and observational data that may reflect patient condition changes earlier than other physiological values^12,46,47^ and are information-rich^13,48^, but often not well understood outside of nursing.

CONCERN EWS was developed by an interdisciplinary team that employed rigorous methods for translating a predictive model to the clinical setting, including nurse and prescribing provider input on healthcare process effects and real-time data availability, integration with existing clinical workflows, transparency and explainability to gain trust^12,49^, as well as robust evaluation for external validity^50^ and model fairness. This comprehensive approach resulted in 42-hour greater lead time than other EWSs^8^ which allows for identification of deteriorating patients several clinical shifts before an event resulting in greater time and opportunity for clinically meaningful interventions. By leveraging nursing surveillance patterns, rather than physiological measures^12,47^, CONCERN EWS overcomes the limitations of physiological measurements^8^ and leverages an existing expert knowledge base: nurses’ autonomous decisions to take greater action and document more observations or notes than mandated by policy in direct response to concerning changes in a patient’s clinical state.^12,13^ Mortality increases when care escalation is delayed.^51^ Therefore, transparency and explainability, achieved by displaying the score and factors that drove the prediction, may overcome the dissonance between nurses’ identification of patient risk based on subtle clinical indicators and prescribing providers’ expectations of changes in physiological data, thereby prompting earlier decisions to escalate medical intervention. We anticipate several future clinical applications to our CONCERN EWS approach, including expansion to other clinical practice patterns and patient populations within and beyond the hospital setting.

A major purpose of hospitalization is to provide continuous, “around-the-clock” nursing care that would not be feasible in an outpatient setting, yet health care systems and clinicians are undergoing many challenges today. Despite the well-documented poor staffing, increased patient acuity, and limited hospital resources rampant across the globe during the time we conducted our study (2020-2022)^1,52^, CONCERN EWS still improved outcomes over usual-care across both study sites. The predictors in CONCERN EWS are based on the documented decisions that a nurse makes about how and when to provide nursing care. As such, CONCERN EWS is a novel approach to measuring nursing value and capitalizing on nursing expertise, which is critically needed amidst this unprecedented time of clinician burnout and turnover.^52^

### 4.1 Limitations

The reported findings have important limitations. First, the trial was complicated by disruptions at the sites due to the COVID-19 pandemic, which forced a 1-year postponement at one of our sites due to implementation delays. Second, the trial was conducted at 2 health systems located in urban areas within the northeastern United States, and findings may not be generalizable to hospitals in different settings, especially in other countries which may have different nursing practices.

## Conclusion

Our study demonstrates that nursing surveillance patterns are a valuable signal to predict deterioration of hospitalized patients. A hospital-wide EWS based on nursing surveillance patterns resulted in a 35.6% decreased risk of in-hospital mortality, 11.2% decreased LOS, 7.5% decreased risk of sepsis, and 24.9% increased risk of unanticipated ICU transfers, when integrated into the care team’s EHR workflow compared with usual-care. Continuing CONCERN EWS work includes spreading to additional sites and countries, implementation science evaluations, and expansion of the predictive model to other hospital units (e.g., emergency department) and inpatient populations (e.g., pediatrics).

## Author Contributions

Drs. Rossetti and Cato had full access to all the data in the study and take responsibility for the integrity of the data and the accuracy of the data analysis. More than one author (Cato, Rossetti, Lowenthal, Jia, Tran, Withall, Lee) have directly accessed and verified the underlying data reported in the manuscript.

*Concept:* Rossetti, Cato

*System development and study design:* Rossetti, Cato, Dykes, Knaplund, Cho, Albers, Lowenthal, Bakken, Kang, Chang, Zhou, Withall, Jia, Liu, Schwartz-Dillard

*Acquisition, analysis, or interpretation of data:* Rossetti, Cato, Dykes, Knaplund, Cho, Albers, Lowenthal, Bakken, Withall, Jia, Liu, Lee, Daramola, Tran, Bokhari, Thate

*Drafting of the manuscript:* Rossetti, Lee, Cato, Dykes

*Critical review of the manuscript for important intellectual content:* All authors.

*Statistical analysis:* Jia, Cato, Albers

*Obtained funding:* Rossetti, Cato

*Administrative, technical, or material support:* Rossetti, Cato, Dykes, Lee, Daramola, Liu

*Supervision:* Rossetti, Cato, Dykes, Bakken

## Conflict of Interest Disclosures

Dr. Bates reports grants and personal fees from EarlySense, personal fees from CDI Negev, equity from ValeraHealth, equity from Clew, equity from MDClone, personal fees and equity from AESOP, personal fees and equity from FeelBetter, personal fees and equity from Guided Clinical Solutions and grants from IBM Watson Health, outside the submitted work.

## Funding/Support

This study was funded by the National Institute of Nursing Research (NINR 1R01NR016941, COmmunicating Narrative Concerns Entered by RNs (CONCERN): Clinical Decision Support Communication for Risky Patient States) and Reducing Health Disparities Through Informatics (T32NR007969). The funder of the study had no role in study design, data collection, data analysis, data interpretation, or writing of the report. The content is solely the responsibility of the authors and does not necessarily represent the official views of the National Institutes of Health.

## Data Sharing Statement

Individual participant data including data dictionaries and study protocol will be made available. Specifically, de-identified individual shift level and encounter level data for primary and secondary outcomes in this trial will be made available after de-identification. Data will be made available within 6 months following article publication with no anticipated end date. We will make the data available to investigators whose proposed use of the data has been approved by an independent review board. Data will be available through our study website https://www.dbmi.columbia.edu/concern-study/. To gain access, data requestors will need to sign a data access agreement. We will share statistical analysis code upon request and additional information, including study protocol details, is available in our online supplement. The CONCERN EWS algorithm is considered intellectual property that will not be shared but is described in our online supplement.

## Additional Contributions

We thank all the nurses, prescribing providers, and patients who participated in this study. We also acknowledge Bonnie Westra, RN, PhD, for serving on our Advisory Board and the American Nurses Foundation Reimagining Nursing Initiative funding to spread and evaluate the implementation of CONCERN EWS to additional study sites. We thank Carolyn Stillwell for her contribution to the design of tables and figures and Lorene Schweig for editorial review.

## Supporting information

Online Supplement CONCERN Trial

## Notes

### Clinical Trial

NCT03911687

### Clinical Protocols

https://pubmed.ncbi.nlm.nih.gov/34889766/

### Author Declarations

Institutional review boards of Columbia University and Brigham and Women's Hospital gave ethical approval for this work.

