## Supplementary material for "Multisite Pragmatic Cluster-Randomized Controlled Trial of the CONCERN Early Warning System": Online Supplement CONCERN Trial

### Table of Contents


| Content | Page |
|---|---|
| **eTable1.** Count of Study Units by Type and Intervention versus Usual Care Group Across Study Sites | 2 |
| **eTable2.** Performance Metrics for CONCERN Predictive Model | 3 |
| **eTable3.** CONCERN Model Factors and Features | 4 |
| **eTable4.** Statistical Power Analysis | 5 |
| **eTable5.** Characteristics of Patients with Hospital Encounters During the Trial | 6 |
| **eTable6.** Top 10 Admission and Principal Diagnoses Across Study Sites | 7 |
| **eFigure1.** CONCERN Predictive Model Conceptual Modeling Approach | 8 |
| **eMethods1.** Summary of Changes from Original to Final Study Protocol and Analysis Plans | 9 |
| **eMethods2.** Detailed Description of CONCERN Early Warning System (EWS) Intervention | 10 |
| **eTable7.** Reporting of trial findings according to the CONSORT guidelines and extensions for AI, cluster-randomized trials, and pragmatic trials | 11-20 |
| **References** | 21 |

**eTable1. Count of Study Units by Type and Intervention versus Usual Care Group Across Study Sites**

| | Site A | | Site B | | |
|---|---|---|---|---|---|
| Unit Type | CONCERN Intervention | Usual Care | CONCERN Intervention | Usual Care | Totals |
| Acute Care Unit (ACU) | 19 | 17 | 9 | 8 | 53 |
| Intensive Care Unit (ICU) | 5 | 7 | 4 | 5 | 21 |
| Totals | 24 | 24 | 13 | 13 | 74 |

**eTable2. Performance Metrics for CONCERN Predictive Model[a]**

| Setting | Accuracy | Precision | Recall | Log Loss | AUC |
|---|---|---|---|---|---|
| ICU | 0.970938 | 0.431373 | 0.594595 | 0.073695 | 0.934683 |
| ACU | 0.973341 | 0.813559 | 0.643935 | 0.089369 | 0.955982 |

[a]Multinomial Gradient Boosted Machine (GBM) model built on random 12-hour time slices to predict (over the next 24 hours) whether a patient is discharged, will still be in the hospital or has a hospital event (in-hospital mortality, cardiopulmonary arrest, sepsis, unanticipated ICU transfer). Modeling was trained on 70% of the dataset, with 30% used for 10-fold cross-validation. Average performance reported for ensemble models.[1]

**eTable3. CONCERN Model Factors and Features**

| | |
|---|---|
| **Nursing Note Content** | |
| | Abdominal pain* |
| | Abnormal heart rhythm* |
| | Abnormal mental state* |
| | Abnormal rate, rhythm, depth and effort of respirations* |
| | Abnormal temperature* |
| | Back pain* |
| | Chest pain* |
| | Communication problem* |
| | Diagnosis related with infection* |
| | Deficit of circulation* |
| | Fall risk* |
| | Fluid volume alteration* |
| | General concern* |
| | Headache* |
| | Improper renal function* |
| | Medication related with infection* |
| | Monitoring* |
| | Mood disorder* |
| | Musculoskeletal pain* |
| | Pain level* |
| | Violence gesture* |
| **Vital Sign Frequency** | |
| | Heart rate measurement* |
| | Respiratory rate measurement* |
| | Blood pressure measurement* |
| | Temperature measurement* |
| | $SpO_2$ measurement* |
| | All 5 vital measurements taken at same time* |
| | Only 1 vital measurement taken* |
| **Nursing Note Frequency** | |
| | Nursing note written* |
| **Vital Sign Comment Frequency** | |
| | Heart rate comment* |
| | Respiratory rate comment* |
| | Blood pressure comment* |
| | Temperature comment* |
| | $SpO_2$ comment* |
| **Medication Administration** | |
| | PRN medication administered* |
| | Scheduled medication withheld* |
| * Feature is aggregated over the past 12 hours | |

**eTable4. Statistical Power Analysis**

| | |
|---|---|
| Original analysis plan | • Generalized linear mixed models for the comparison of mortality rates between those randomized to CONCERN and usual care |
| Parameters | • 2-sided tests with alpha = 0.05<br>• Expected sample:<br>○ 2,000 total admissions per month<br>○ 50,000 total inpatient days based on average occupancy<br>○ Mean mortality rate of 37.5 deaths per 10,000 inpatient days (ranging from 11.9 to 48.8 across our study sites) |
| Estimated effect size | • 80% statistical power to detect a difference of less than 1% relative difference in mortality rates |

**eTable5. Characteristics of Patients with Hospital Encounters During the Trial (N=60,893)**

| | Site A | | Site B | |
|---|---|---|---|---|
| | CONCERN Intervention<br><br>N = 16,838 | Usual Care<br><br>N = 15,630 | CONCERN Intervention<br><br>N = 16,186 | Usual Care<br><br>N = 12,239 |
| Age | 63.03 ± 16.86 | 63.72 ± 16.67 | 62.19 ± 18.26 | 63.60 ± 17.62 |
| Male Sex – no. (%) | 8,276 (49.15) | 7,543 (48.26) | 7,780 (48.07) | 6,423 (52.48) |
| Race – no. (%) | | | | |
| White | 13,483 (80.07) | 12,327 (78.87) | 5,440 (33.61) | 3,951 (32.28) |
| Black | 1,564 (9.29) | 1,540 (9.85) | 3,314 (20.47) | 2,684 (21.93) |
| Asian | 455 (2.70) | 467 (2.99) | 248 (1.53) | 167 (1.36) |
| Other or missing | 1,336 (7.93) | 1,296 (8.29) | 7,184 (44.38) | 5,437 (44.42) |
| Ethnic group – no. (%) | | | | |
| Not Hispanic or Latino | 15,298 (90.85) | 14,200 (90.85) | 8,320 (51.40) | 6,104 (49.87) |
| Hispanic or Latino | 1,172 (6.96) | 1,151 (7.36) | 6,270 (38.74) | 4,752 (38.83) |
| Unknown or not reported | 368 (2.19) | 279 (1.79) | 1,596 (9.86) | 1,383 (11.30) |
| Primary Language English | 15,662 (93.02) | 14,378 (91.99) | 11,051 (68.28) | 8,206 (67.05) |
| Charlson Comorbidity Index | 3.56 ± 3.34 | 4.01 ± 3.53 | 2.64 ± 2.89 | 2.87 ± 2.97 |
| Discharge Disposition[$] | | | | |
| Home | 13,667 | 12,860 | 13,106 | 9,346 |
| Other | 3,171 | 2,770 | 3,080 | 2,893 |

± *Plus–minus values are mean* ±SD
*Race and ethnic group were reported in the EHR
[$]Discharge Disposition: Home includes home with services; Other includes any disposition not to home

**eTable6. Top 10 Admission and Principal Diagnoses[a] Across Study Sites**

| Rank | Site A Top 10 Admission Diagnoses | Site B Top 10 Admission Diagnoses |
|---|---|---|
| 1 | Shortness of breath | Shortness of breath |
| 2 | Unspecified abdominal pain | Unspecified abdominal pain |
| 3 | Chest pain, unspecified | Chest pain, unspecified |
| 4 | Fever, unspecified | Sepsis, unspecified organism |
| 5 | Altered mental status, unspecified | Altered mental status, unspecified |
| 6 | Weakness | COVID-19 |
| 7 | COVID-19 | Syncope and collapse |
| 8 | Morbid (severe) obesity due to excess calories | Weakness |
| 9 | Solitary pulmonary nodule | Nonrheumatic aortic (valve) stenosis |
| 10 | Nausea with vomiting, unspecified | Fever, unspecified |
| **Rank** | **Site A Top 10 Principal Diagnoses** | **Site B Top 10 Principal Diagnoses** |
| 1 | COVID-19 | Sepsis, unspecified organism |
| 2 | Hypertensive heart and chronic kidney disease with heart failure and stage 1 through stage 4 chronic kidney disease, or unspecified chronic kidney disease | COVID-19 |
| 3 | Hypertensive heart disease with heart failure | Hypertensive heart and chronic kidney disease with heart failure and stage 1 through stage 4 chronic kidney disease, or unspecified chronic kidney disease |
| 4 | Non-ST elevation (NSTEMI) myocardial infarction | Hypertensive heart disease with heart failure |
| 5 | Spinal stenosis, lumbar region with neurogenic claudication | Nonrheumatic aortic (valve) stenosis |
| 6 | Morbid (severe) obesity due to excess calories | Non-ST elevation (NSTEMI) myocardial infarction |
| 7 | Acute kidney failure, unspecified | Morbid (severe) obesity due to excess calories |
| 8 | Sepsis, unspecified organism | Atherosclerotic heart disease of native coronary artery without angina pectoris |
| 9 | Nonrheumatic aortic (valve) stenosis | Acute kidney failure, unspecified |
| 10 | Spinal stenosis, cervical region | Hypertensive heart and chronic kidney disease with heart failure and with stage 5 chronic kidney disease, or end stage renal disease |

[a]ICD10 Codes; Gray shading indicates present in Top 10 list for both study sites

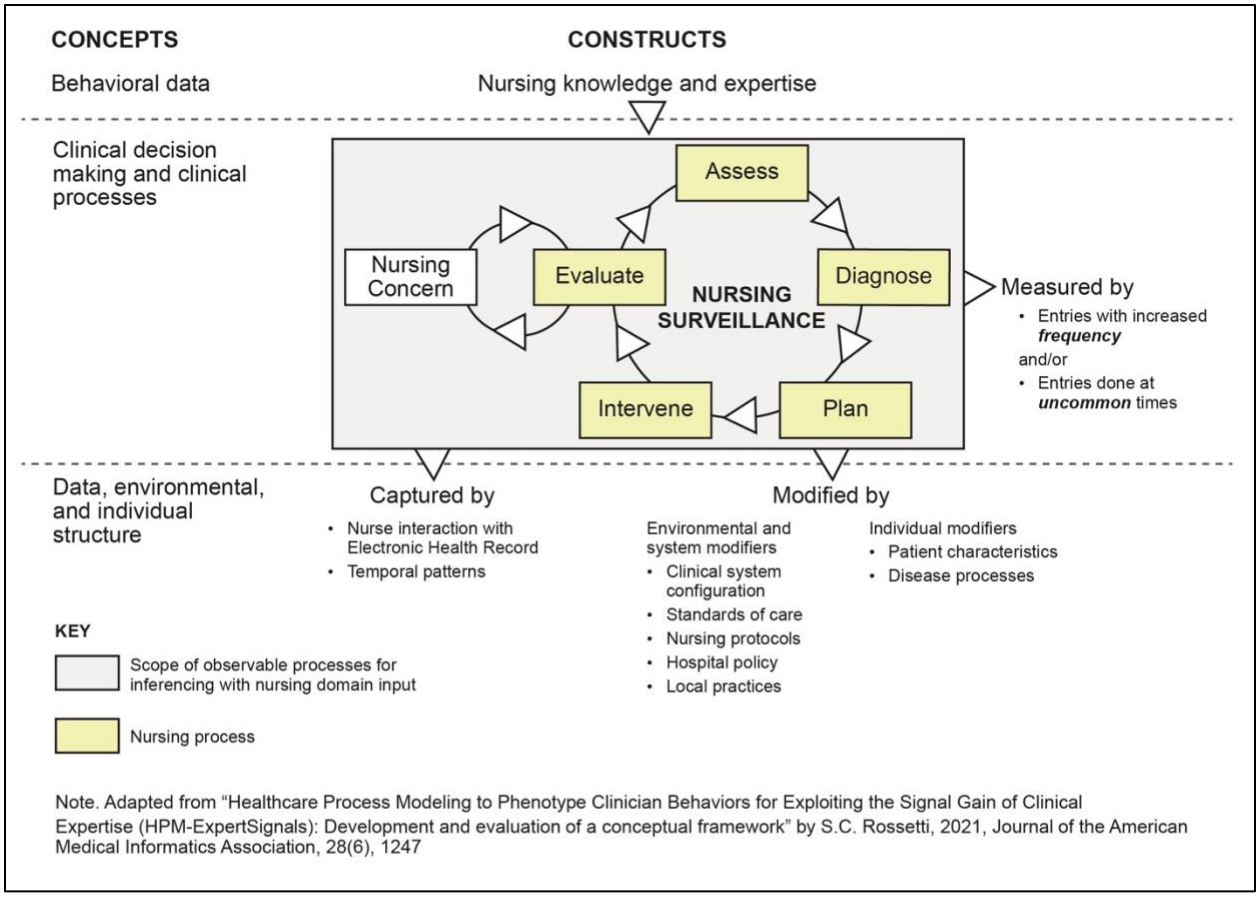


**eFigure1. CONCERN Predictive Model Conceptual Modeling Approach**

### eMethods1: Summary of Changes from Original to Final Study Protocol and Analysis Plans

Our original study protocol is published here[3]. Below we describe all changes from the original protocol to our final protocol and rationale for those changes. We note that the trial's two study sites, primary and secondary outcomes, and enrollment eligibility remained the same as originally proposed.

**Study Design**

We originally planned to use generalized linear mixed models for an interrupted time series analysis with six-month pre-intervention data at the clinical unit level of analysis. However, and as noted in the main manuscript, since a significant proportion of patients moved across different units during their hospital stay, it was not possible. Instead, in our final protocol we analyzed all outcomes at the individual patient level.

The original study design was described as a multiple time-series intervention. The final study design was a cluster-randomized pragmatic controlled clinical trial. All study design changes were made prior to randomization and prior to the beginning of the trial in October 2020. The original protocol design stated that non-equivalent control units (usual care) would be used for comparison. Instead, we selected to use a cluster-randomized approach for the random assignment of acute care units and intensive care units (ICUs) to intervention or usual care groups prior to the start of the trial and stratified by site. Based on this final study design, the originally proposed six-months of pre-intervention data was not needed. In selection of our final study design, we considered several study design approaches, including cross-over trial design, and their ramifications, including ethical and legal considerations for a clinical decision support intervention, and determined a cluster-randomized pragmatic controlled clinical trial was the most appropriate and rigorous design.[4] Informatics interventions in the hospital setting, such as the CONCERN EWS system, are best randomized at the clinical unit level to avoid risk of cross-over both within patient assignments and across care team members if randomized at the individual patient or clinician level.[4] The nature of the intervention being clinical decision support precluded blinding; however, we used objective primary and secondary outcomes queried directly from the EHR with blinded validation through chart reviews.[4]

The original protocol allowed for continuous refinement of our machine learning algorithm, pushing out a new version every 3 months and tracking outcomes for each version. This was proposed due to the possibility that nursing documentation patterns may change overtime in response to the use of a novel algorithm that is based on real-time documentation patterns. Our Advisory Board recommended that we perform an interim analysis at our Study Site A (go-live 2020) to confirm model performance before our Study site B go-live in 2021 to inform if model versioning was needed or appropriate. Based on the interim analysis we concluded that no updates were needed to the model, therefore the same version of the model was used during the entirety of the trial at both study sites.

We note that our trial is described as a pragmatic trial. Our original protocol written in 2017 was prior to Study Site B changing electronic health records in February of 2020 and the COVID-19 pandemic starting in spring 2020. Both events influenced unavoidable delays in the trial at Study Site B. Originally our trial was planned to be conducted simultaneously at both study sites. Instead, we conducted a pragmatic, but rigorous trial at Study Site A in 2020-2021 and Study Site B 2021-2022.

**Statistical Analysis Plan**

Our final trial sample size was based on our original power analysis, which is provided in in the main manuscript. Given the same version of the model was tested during the entirety of the trial a multiple time-series intervention analysis was no longer appropriate.

At an early stage of the interim analysis, we noted that it was impossible to perform unit level analysis for interrupted time series models and made modifications to our statistical plan at that point. As described above, we originally planned to use generalized linear mixed models for an interrupted time series analysis at the unit level. However, a significant proportion of patients moved across different units during their hospital stay. Therefore, all originally proposed unit level analyses described in the original statistical analysis plan above were inappropriate. Instead, we analyzed all outcomes at the individual patient level. While we did remove the unit level analysis (interrupted time series) from the final analyses, the rest of our final analytical plan is not different from that in the original proposal. The original analysis plan specified use of generalized linear mixed model for LOS (Poisson models) and readmission (logit link function) and survival models for other outcomes, which is consistent with our final analytical plan. Statistical analyses were performed using SAS 9.4 Copyright (c) 2002-2012 by SAS Institute Inc., Cary, NC, USA.

### eMethods2: Detailed Description of CONCERN Early Warning System (EWS) Intervention

**CONCERN EWS Prediction: Predictive Model Output as Early Warning Score of Patient Risk**

Our risk prediction model (version 1.5) is nonlinear, with time-varying variables (e.g., vital signs recorded at variable times within a clinical shift). Every hour, the CONCERN EWS processes EHR data from the past 24 hours through the ensemble models that best fit the patient characteristics, calculating a risk score (green, yellow, or red). See eFigure1 for conceptual modeling approach and eTable3 for model factors and features. CONCERN EWS' conceptual modeling approach and model factors can be summarized as reflecting patterns of increased nurses' surveillance above and beyond the standard of care and resultant nursing interventions consistent with an observed change in the patient's clinical state. Increased nurses' surveillance in our model, for example, may be measured when a nurse assesses vital signs every 2 hours for a specific patient displaying subtle changes even though the clinical unit policy only requires assessment every 6 hours. As an example of a nursing intervention consistent with observed but subtle changes in the patient's clinical state, a nurse may decide not to administer a scheduled medication, such as metoprolol, because the patient's heart rate is hovering around 60 beats per minute and their usual baseline is around 90 beats per minute. While medications are ordered by a prescribing provider in the hospital setting, the administration is completed by a nurse and requires the nurse to assess the appropriateness of that medication given the patient's condition prior to administration. Finally, our time-varying modeling approach also considers if nursing assessments or interventions are performed at non-common times, based on our data-driven analyses. The CONCERN EWS includes a monitoring function to check that all variables are available to the model and that the volume of data for each variable is consistent with historical volumes, with automated emailed notification of issues. The CONCERN EWS has robust error reporting and logging functionalities for each step in our data pipeline, including data inputs, pre-processing, prediction generation, and CONCERN score writing to the EHR. A system performance error was defined as the automated prediction score not being generated every hour for eligible patients. The few times that automated prediction did not occur during our trial were due to a web service error. The research team was alerted and able to manually trigger the prediction processes for all patients.

Approaches to evaluate the predictive accuracy of a nonlinear, time-varying model must consider the clinical problem, data inputs, and model features. See eTable2 for Performance Metrics for CONCERN Predictive Model. Through experimentation on retrospective data sets of hospitalized patients' encounters in the EHR, we refined our machine learning modeling approach (see eFigure1), which is focused on temporal patterns of nursing observations, using ensemble learning to identify the best performing models to predict clinical deterioration in the next 24 hours, for a patient's specific clinical situation. For our model building, patient deterioration was defined as the first occurrence of unanticipated transfer to the ICU, sepsis, mortality, or cardiopulmonary arrest. For example, ensemble learning allowed us to identify the best model for a patient who has been in an ICU for 5 hours on a Friday night and to make our EWS transparent and explainable. We are not aware of any other decision-support implementations of EWSs that use a similar multi-factorial, advanced computational, *and* explainable modeling approach.[2]

**CONCERN EWS Display: Clinical Decision Support in the EHR to Increase Team Situational Awareness of Patient Risk**

Clinical decision support functionality was developed based on substantial user-centered design sessions and simulation evaluations and was integrated into the EHR using the FHIR (Fast Healthcare Interoperability Resources) standard for exchanging healthcare information electronically.[2] The Clinical Decision Support displayed the early warning score (green, yellow, red) in the EHR patient list for all nurses and prescribing providers (physicians and advanced practice providers) on the clinical unit. Displaying the score in the EHR patient list meant that the score was visible on nurses' and prescribing providers' log-in screen and part of the typical EHR workflow when selecting a new patient chart to view. Nurses and prescribing providers could double-click the score icon linked to a screen with detailed information about the prediction (Figure 1)[3], including the factors that determined a patient's score, a score trendline adjustable up to 72 hours, and a scale displaying the patient's score relative to other hospitalized patients. Users could also click on the factors that determined the score to view the clinical data associated with that factor. The factors are comprised of one or more granular CONCERN EWS features (e.g., vital sign frequency factor includes measurement patterns related to each type of vital sign).

**CONCERN EWS Intervention Implementation Activities**

As part of the intervention to ensure hospital-level support and end-user awareness of the score and information displayed within the EHR, we created robust stakeholder engagement and end-user training processes. As with most information technology implementations in the clinical setting, early and frequent engagement sessions were held with stakeholders at all levels within the hospital, including nurse and physician leadership, registered nurses, prescribing providers, clinical champions, analysts, technicians, and other hospital leaders. We also created training materials and training processes for super-users and end-users (nurses and prescribing providers) that conveyed information about the EWS score and its display within the EHR

**eTable7. Reporting of trial findings according to the CONSORT guidelines and extensions for AI, cluster-randomized trials, and pragmatic trials**

| Section/Topic | Item No. | CONSORT 2010 item | Extension for AI studies | Extension for cluster randomized trials | Extension for pragmatic trials | Page No. (Section) |
|---|---|---|---|---|---|---|
| **Title and abstract** | | | | | | |
| Title and abstract | 1a | Identification as a randomised trial in the title | CONSORT-AI 1a,b Elaboration:<br><br>Indicate that the intervention involves artificial intelligence/machine learning in the title and/or abstract and specify the type of model | Identification as a cluster randomised trial in the title | | Pg.1 (Title page), Pg. 3-4 (Abstract) |
| | 1b | Structured summary of trial design, methods, results, and conclusions (for specific guidance see CONSORT for abstracts) | State the intended use of the AI intervention within the trial in the title and/or abstract | See extension for CONSORT-abstracts | | Pg.3-4 (Abstract) |
| **Introduction** | | | | | | |
| Background and objectives | 2a | Scientific background and explanation of rationale | CONSORT-AI 2a (i) Extension<br><br>Explain the intended use of the AI intervention in the context of the clinical pathway, including its purpose and its intended users (for example, healthcare professionals, patients, public). | Rationale for using a cluster design | Describe the health or health service problem that the intervention is intended to address and other interventions that may commonly be aimed at this problem | Pg.5-6 (1.Introduction)<br><br>Online Supplement eMethods1 |

| | | | | | | |
|---|---|---|---|---|---|---|
| | 2b | Specific objectives or hypotheses | | Whether objectives pertain to the cluster level, the individual participant level, or both | | Pg.6 (1.Introduction) |
| **Methods** | | | | | | |
| Trial design | 3a | Description of trial design (such as parallel, factorial) including allocation ratio | | Definition of clusters and description of how the design features apply to the clusters | | Pg. 6-7 (2.1 Study Design, Trial Sites, and Randomization) |
| | 3b | Important changes to methods after trial commencement (such as eligibility criteria), with reasons | | | | Pg. 6-7 (2.1 Study Design, Trial Sites, and Randomization)<br><br>Online Supplement eMethods1 |
| Participants | 4a | Eligibility criteria for participants | CONSORT-AI 4a (i) Elaboration : State the inclusion and exclusion criteria at the level of participants.<br><br>CONSORT-AI 4a (ii) Extension<br>State the inclusion and exclusion criteria at the level of the input data. | Eligibility criteria for clusters | | Pg. 6-8 (2.1 Study Design, Trial Sites, and Randomization; 2.2 Trial Participants and Outcomes) |
| | 4b | Settings and locations where the data were collected | CONSORT-AI 4b Extension<br>Describe how the AI intervention was integrated into the trial setting, including any onsite or offsite requirements. | | | Pg.6-7 (2.1 Study Design, Trial Sites, and Randomization) |
| Interventions | 5 | The interventions for each group with sufficient details to | CONSORT-AI 5 (i) Extension | Whether interventions pertain to the cluster level, | Describe extra resources added to (or resources | Pg. 6-8 (2.1 Study Design, Trial Sites, and Randomization, Figure |

| | | | | | | |
|---|---|---|---|---|---|---|
| | | allow replication, including how and when they were actually administered | State which version of the AI algorithm was used.<br>CONSORT-AI 5 (ii) Extension<br>Describe how the input data were acquired and selected for the AI intervention.<br><br>CONSORT-AI 5 (iii) Extension<br>Describe how poor quality or unavailable input data were assessed and handled.<br>CONSORT-AI 5 (iv) Extension<br>Specify whether there was human–AI interaction in the handling of the input data, and what level of expertise was required of users.<br><br>CONSORT-AI 5 (v) Extension<br>Specify the output of the AI intervention<br><br>CONSORT-AI 5 (vi) Extension<br>Explain how the AI intervention's outputs contributed to decision-making or other elements of clinical practice. | the individual, participant level, or both | removed from) usual settings in order to implement intervention. Indicate if efforts were made to standardise the intervention or if the intervention and its delivery were allowed to vary between participants, practitioners, or study sites<br><br>Describe the comparator in similar detail to the intervention | 1, 2.3 CONCERN EWS Intervention)<br>Online Supplement eFigure 1, eTable2, eTable 3, eMethods2 |

| | | | | | | |
|---|---|---|---|---|---|---|
| Outcomes | 6a | Completely defined pre-specified primary and secondary outcome measures, including how and when they were assessed | | Whether outcome measures pertain to the cluster level, the individual participant level, or both | Explain why the chosen outcomes and, when relevant, the length of follow-up are considered important to those who will use the results of the trial | Pg.7-11 (2.2 Trial Participants and Outcomes, Table 1, 2.4 Statistical Analysis, 2.4.1. LOS Outcome, 2.4.2. Death and Secondary In-Hospital Event Outcomes, 2.4.3. Transfers Between Intervention-and Usual Care Units, Table 3 ) |
| | 6b | Any changes to trial outcomes after the trial commenced, with reasons | | | | Online Supplement eMethods1 |
| Sample size | 7a | How sample size was determined | | Method of calculation, number of clusters(s) (and whether equal or unequal cluster sizes are assumed), cluster size, a coefficient of intracluster correlation (ICC or k), and an indication of its uncertainty | If calculated using the smallest difference considered important by the target decision maker audience (the minimally important difference) then report where this difference was obtained | Pg. 9 (2.4 Statistical Analysis)<br><br>Online Supplement eTable4 |
| | 7b | When applicable, explanation of any interim analyses and stopping guidelines | | | | N/A |
| **Randomization** | | | | | | |
| Sequence generation | 8a | Method used to generate the | | | | Pg. 6-7 (2.1 Study Design, Trial Sites, and Randomization) |

| | | | | | | |
|---|---|---|---|---|---|---|
| | | random allocation sequence | | | | |
| | 8b | Type of randomisation; details of any restriction (such as blocking and block size) | | Details of stratification or matching if used | | |
| Allocation concealment mechanism | 9 | Mechanism used to implement the random allocation sequence (such as sequentially numbered containers), describing any steps taken to conceal the sequence until interventions were assigned | | Specification that allocation was based on clusters rather than individuals and whether allocation concealment (if any) was at the cluster level, the individual participant level, or both | | Pg. 6-7 (2.1 Study Design, Trial Sites, and Randomization) |
| Implementation | 10 | Who generated the random allocation sequence, who enrolled participants, and who assigned participants to interventions | | Replaced by 10a, 10b, and 10c | | Pg. 7 (2.1 Study Design, Trial Sites, and Randomization) |
| | 10a | | | Who generated the random allocation sequence, who enrolled clusters, and who assigned clusters to interventions | | Pg. 7 (2.1 Study Design, Trial Sites, and Randomization) |
| | 10b | | | Mechanism by which individual participants were | | Pg.7 (2.1 Study Design, Trial Sites, and Randomization) |

| | | | | | | |
|---|---|---|---|---|---|---|
| | | | | included in clusters for the purposes of the trial (such as complete enumeration, random sampling) | | |
| | 10c | | | From whom consent was sought (representatives of the cluster, or individual cluster members, or both) and whether consent was sought before or after randomisation | | Pg.7 (2.1 Study Design, Trial Sites, and Randomization) |
| Blinding | 11a | If done, who was blinded after assignment to interventions (for example, participants, care providers, those assessing outcomes) and how | | | If blinding was not done, or was not possible, explain why | Online Supplement eMethods1 |
| | 11b | If relevant, description of the similarity of interventions | | | | N/A |
| Statistical methods | 12a | Statistical methods used to compare groups for primary and secondary outcomes | | How clustering was taken into account | | Pg. 9-11 (2.4 statistical analysis)<br><br>Online Supplement eMethods1 |

| | | | | | | |
|---|---|---|---|---|---|---|
| | 12b | Methods for additional analyses, such as subgroup analyses and adjusted analyses | | | | N/A |
| **Results** | | | | | | |
| Participant flow (a diagram is strongly recommended) | 13a | For each group, the numbers of participants who were randomly assigned, received intended treatment, and were analysed for the primary outcome | | For each group, the numbers of clusters that were randomly assigned, received intended treatment, and were analysed for the primary outcome | The number of participants or units approached to take part in the trial, the number which were eligible, and reasons for non-participation should be reported | Pg. 11 (3.1 Trial Participants and Hospital Encounters, Figure 2)<br><br>Online Supplement eTable1 |
| | 13b | For each group, losses, and exclusions after randomisation, together with reasons | | For each group, losses and exclusions for both clusters and individual cluster members | | Pg. 11 (3.1 Trial Participants and Hospital Encounters, Figure 2) |
| Recruitment | 14a | Dates defining the periods of recruitment and follow-up | | | | Pg. 7 (2.1 Study Design, Trial Sites, and Randomization)<br><br>Online Supplement eMethods1 |
| | 14b | Why the trial ended or was stopped | | | | N/A |
| Baseline data | 15 | A table showing baseline demographic and clinical characteristics for each group | | Baseline characteristics for the individual and cluster levels as applicable for each group | | Pg. 11 (3.1 Trial Participants and Hospital Encounters, Table 2)<br><br>Online Supplement eTable1, eTable 5, eTable 6 |

| | | | | | | |
|---|---|---|---|---|---|---|
| Numbers analyzed | 16 | For each group, number of participants (denominator) included in each analysis and whether the analysis was by original assigned groups | | For each group, number of clusters included in each analysis | | Pg. 11 (3.1 Trial Participants and Hospital Encounters) |
| Outcomes and estimation | 17a | For each primary and secondary outcome, results for each group, and the estimated effect size and its precision (such as 95% confidence interval) | | Results at the individual or cluster level as applicable and a coefficient of intracluster correlation (ICC or k) for each primary outcome | | Pg. 12 (3.2 Primary Outcomes, 3.3 Secondary Outcomes, Table 3) |
| | 17b | For binary outcomes, presentation of both absolute and relative effect sizes is recommended | | | | Pg. 12 (3.2 Primary Outcomes, 3.3 Secondary Outcomes, Table 3) |
| Ancillary analyses | 18 | Results of any other analyses performed, including subgroup analyses and adjusted analyses, distinguishing pre-specified from exploratory | | | | N/A |
| Harms | 19 | All important harms or unintended effects in each group (for specific guidance see CONSORT for harms) | CONSORT-AI 19 Extension<br>Describe results of any analysis of performance errors and how errors were identified, where applicable. If no such | | | Online Supplement eMethods2 |

| | | | | | | |
|---|---|---|---|---|---|---|
| | | | analysis was planned or done, justify why not. | | | |
| **Discussion** | | | | | | |
| Limitations | 20 | Trial limitations, addressing sources of potential bias, imprecision, and, if relevant, multiplicity of analyses | | | | Pg.15 (4.1 Limitations) |
| Generalizability | 21 | Generalisability (external validity, applicability) of the trial findings | | Generalisability to clusters and/or individual participants (as relevant) | Describe key aspects of the setting which determined the trial results.<br><br>Discuss possible differences in other settings where clinical traditions, health service organisation, staffing, or resources may vary from those of the trial | Pg.15 (4.1 Limitations) |
| Interpretation | 22 | Interpretation consistent with results, balancing benefits, and harms, and considering other relevant evidence | | | | Pg. 12-15 (4. Discussion) |
| **Other Information** | | | | | | |
| Registration | 23 | Registration number and name of trial registry | | | | Pg. 1(Title page) |

| | | | | | | |
|---|---|---|---|---|---|---|
| Protocol | 24 | Where the full trial protocol can be accessed, if available | | | | Rossetti SC, Dykes PC, Knaplund C, et al. The Communicating Narrative Concerns Entered by Registered Nurses (CONCERN) Clinical Decision Support Early Warning System: Protocol for a Cluster Randomized Pragmatic Clinical Trial. *JMIR Res Protoc*. 2021;10(12):e30238. doi:10.2196/30238 |
| Funding | 25 | Sources of funding and other support (such as supply of drugs), role of funders | CONSORT-AI 25 Extension<br>State whether and how the AI intervention and/or its code can be accessed, including any restrictions to access or re-use. | | | Pg. 1 (Title page), Pg. 15-16(Funding/Support), Pg. 16 (Data Sharing Statement) |
